# Assessment of the knowledge and attitude of Sri Lankan adults on climate change and its effects on human health

**DOI:** 10.64898/2026.02.07.26345819

**Authors:** I. P. Welgama, U. Muhandiram, T. N. Marikkar, V. Kumarapeli, A. Liyanapathirana

## Abstract

**Introduction:** Climate change is a global adverse phenomenon affecting the health and wellbeing of all humans, and timely awareness can help mitigate these health effects.

**Objective:** To understand the knowledge and attitudes of Sri Lankan adults towards climate change and its effects on human health.

**Methods:** A web based cross-sectional survey was conducted using a structured, pretested, web based, self-administered questionnaire, using a respondent driven sampling technique, among Sri Lankan adults. Data was collected over three months, from 1^st^ September 2022. Responses were automatically stored in a cloud-based database and were imported into a spreadsheet and analysed using MS Excel.

**Results:** Majority of the 118 respondents were young, educated, employed adults from western province, and 56.78% were females. Overall knowledge on climate change was good among 82.20%, while over 90% had a good or favourable knowledge on health effects associated with climate change. Respondents demonstrated a good awareness of climate effects on skin cancer (92.37%), mental illnesses (82.2%) and asthma (82.2%), but were less aware of the effects on diabetes (28.8%), COPD (38.1%) and heart diseases (46.6%), and vector borne diseases such as Malaria (57.6%) and Dengue (61.8%). Over 90% had a good attitude towards the need for climate change mitigation and climate friendly activities being implemented.

**Conclusions:** Urban, educated Sri Lankan adults had a good understanding and awareness on the health effects of climate change, and the importance of mitigating it in relation to its health effects, but further studies are needed to understand the awareness levels of the less educated rural communities.

## Introduction

Climate change is a catastrophic challenge faced by the modern world (1), and profoundly affects all determinants of health, including food security, availability of safe drinking water and a safe habitat. It is defined as a change in the climate, attributed directly or indirectly to human activity, and which alters the composition of the global atmosphere which is in addition to natural climate variability observed over comparable time periods (2-4). This broadly refers to the increased generation and accumulation of greenhouse gases (mainly carbon dioxide, methane and other similar hydrocarbon compounds) within the earth’s atmosphere, altering its composition, and thereby leading to warmer conditions. The outcome is a gross imbalance of all other climate parameters including rainfall, wind and humidity, and deteriorates the atmospheric radiation protective layers such as the ozone layer (4-6). Increased entry of Infra-red radiation, the excessive use of chemicals such as CFC (Chloro-Fluro-Carbons), increased combustion of fossil fuels, mass deforestation, and large-scale farming leading to the excess generation of these greenhouse gases and directly contributes to climate change (5).

The global disease landscape appears to rapidly change with an observable surge in cancer, vector borne diseases, viral respiratory infections, mental illnesses and natural disaster related trauma and malnutrition (6-9). A common risk factor associated with these conditions are the environmental factors, such as exposure to harmful radiation, and gaseous particles, stress related to heat and exhaustion, as well as the impact of natural disasters such as flash floods and hurricanes (8, 9). Despite the massive strides in global healthcare development, little change is observed in the proportions of population affected by lack of safe water and sanitation, lack of adequate food and nutrition, and lack of a safe and stable habitat (10), which are key determinants of health, and the climate impact being identified as a key underlying factor for these drawbacks (11). Climate change has been proven to alter the spread of communicable infections and influence the development and exacerbation of non-communicable diseases (9, 10).

Today, more people are becoming aware of how climate change will affect their health and survival, and how climate change will impact their future generations(12). Thus, climate impact and climate friendly sustainable development have become highly influential decision factors in the current national and global geopolitical arena (12).

In contrast to the situation in most high-income countries, the impact of climate change on the health and survival of populations in Low- or Middle-Income Countries (LMICs) is less well known, especially since socio-economic challenges and political instability exert a more prominent short-term impact on the lives of the people, compared to the longer-term dangers of climate change. Therefore, the knowledge and attitudes of the people with regards to the climate impact on their health could be comparatively less in these settings, exposing the LMIC communities to greater health risks of climate change.

Sri Lanka too has recently experienced major changes in its weather patterns as a result of global climate change^(5)^, but how much are its people aware of its potential health impact needs to be identified. Information in this regard is minimal from Sri Lanka. Considering this information gap, the current study was conducted as an initial community survey to assess the knowledge and attitude of Sri Lankan adults on climate change and its impact on human health.

## Methods

A web based cross sectional survey was conducted within the Sri Lankan cyberspace, using an internet based online, self-administered questionnaire. All persons over the age of 18 years, residing in Sri Lanka for the past 1 year, and were capable of independently understanding and responding to the questionnaire were considered as the study population. The inclusion criteria were the capability of surfing in cyber space with social media applications and E mail. A respondent driven sampling technique was used, with the questionnaire being initially shared with five purposefully selected respondents, whom the investigators were confident would share the questionnaire with their contacts, in the respondent driven manner. This method was adopted with the intention that the questionnaire would be accessible to a larger number of respondents, from all areas of the country. The responses were automatically updated to the database from which a complete MS Excel data sheet was downloadable. Anonymity of responders was maintained as their personal details were not obtained nor recorded. Data was collected over a period of three months from 1^st^ September 2022.

The study tool was a structured, pretested questionnaire. Standard socio demographic questions were included, followed by questions on knowledge and attitudes with regard to climate change, obtained from similar prior studies conducted elsewhere (13-15) and modified considering the local context. Apart from fulfilling the study objectives, it was attempted to utilize this questionnaire as a means of disseminating knowledge on climate change as well among the respondents, and therefore all responses for the questions included were factually correct information. The technical accuracy and comprehension of these questions were reviewed by an expert in climate change, and the face validity and content validity of the questionnaire were established. Thereafter, the questionnaire was refined, pretested, and accurately translated to the local languages.

Climate change was considered to include the rising global temperature and global warming, leading to a meltdown of the polar ice caps and rising sea levels, prolonged periods of unexpected droughts, as well as untimely hurricanes leading to flash floods, and the changes in atmospheric gases leading to acid rains. Furthermore, damage to protective atmospheric layers such as the Ozone layer was also considered, leading to increased radiation risk. Numerous causes leading to climate change were considered to include situations such as rapid deforestation, population growth requiring greater living space and increased waste generation, expansion of livestock farming, increase in industrial development related waste, emission of smoke, steam and toxic fumes into the atmosphere and also the use of numerous harmful agrochemicals (2-6, 16).

The impact of climate change on human health was evidenced by related technical documents and research confirming the associations between climate change and specific disease conditions. Diseases such as skin cancer, lung cancers and leukaemia, respiratory illnesses such as asthma and Chronic Obstructive Pulmonary Disease (COPD), non-communicable diseases such as heart disease, diabetes, and stress and mental health are some diseases of concern (5, 8, 17-20). The increase in incidence of vector borne diseases such as malaria, dengue etc., were well documented (18, 19), while cataract of the eyes have been specially identified as being affected by harmful solar radiation (21). Furthermore, the impending food security crisis due to climate change is already affecting certain regions, leading to a rise in hunger and malnutrition^(4, 18, 19)^.

Regarding implementation of climate friendly interventions, the use of renewable energy sources and minimizing the burning of fossil fuels, green architecture for reducing the energy usage and enhancing the effective use of space for urban development, as well as advancing agriculture through organic farming techniques towards reducing land usage while increasing food production, were all considered important (18, 19). Additionally, efforts on reforestation, and a reduction in harmful emissions of carbon dioxide and other gases, waste segregation and recycling leading to more effective use of natural resources, and minimizing environmental damage, were also considered important actions to mitigate climate change (12, 22).

For knowledge seeking questions, each positive response was given 2 points while not knowing or each negative response earned zero marks. For each section, a total score of 50% or less was considered as “inadequate knowledge” while a score of over 50% was considered as “good knowledge”. Thereafter, to calculate the overall knowledge on the climate impact, the marks for knowledge questions were summed up to obtain a total score. Those who obtained a total summed up score of over 75% were considered to have good overall knowledge while those with a score between 75% and 50% or were considered to have favourable knowledge, and those scoring less than 50% were considered to have poor knowledge.

Similarly, the responses to the questions seeking to assess the attitude were obtained using a Likert scale of 0 to 4, where the response categories were “4 = strongly agree”, “3 = agree”, “2 = neutral / not sure”, “1 = disagree” and “0 = strongly disagree”. Respective marks were awarded based on the response selected by the respondent. Since all questions were related to climate friendly practices, a respondent who agreed with them was considered to have a favourable attitude while those who disagreed or were not sure of the response were considered to have a poor attitude. Following the final summation of response scores, those who had an overall total score of over 75% were considered to have a good attitude, while those scoring between 50% and 75% were considered to have a favourable attitude. Those scoring less than 50% were considered to have a poor attitude.

The findings were presented using frequency tables with the counts and percentages, and the measures of central tendency and dispersion based on the distribution of the sample. The overall knowledge scores, as well as the overall attitude scores, were cross tabulated with sociodemographic variables to identify bivariate associations between these variables and the knowledge and attitude outcomes. Level of significance was defined as 0.05, and Pearson’s Chi Square test was used as the measure of association.

Ethical clearance was obtained from the Ethics review committee of the University of Colombo, Sri Lanka (EC-21-115). Informed consent was obtained prior to the initiation of responses to the online questionnaire, and participation in the study was voluntary.

## Results

Of the 118 respondents, the youngest was 18 years of age while the oldest was 73 years, and most respondents were aged below 40 years. Majority (56. 8%) of the sample were female (Table 1).

**Table 1.**
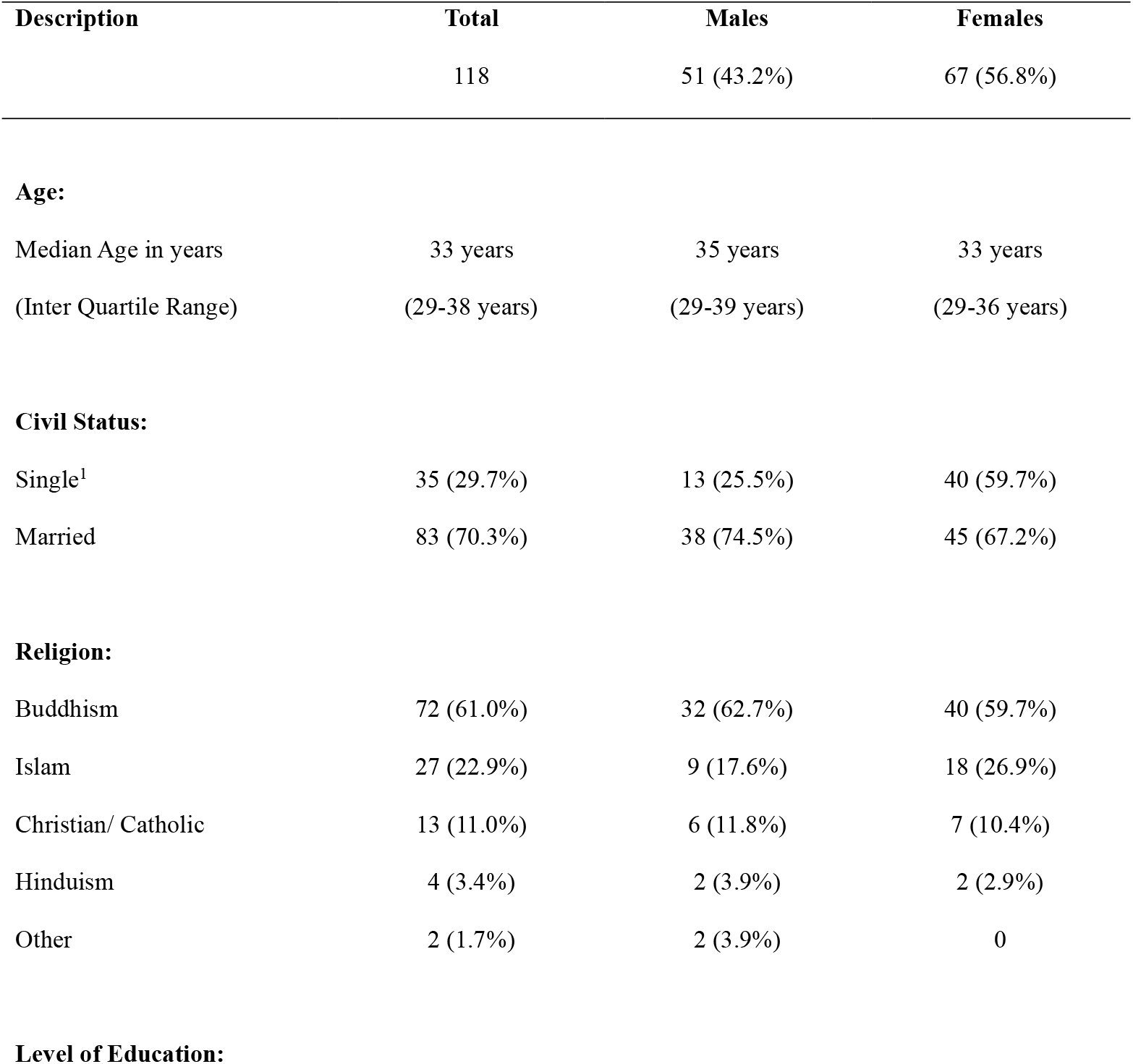
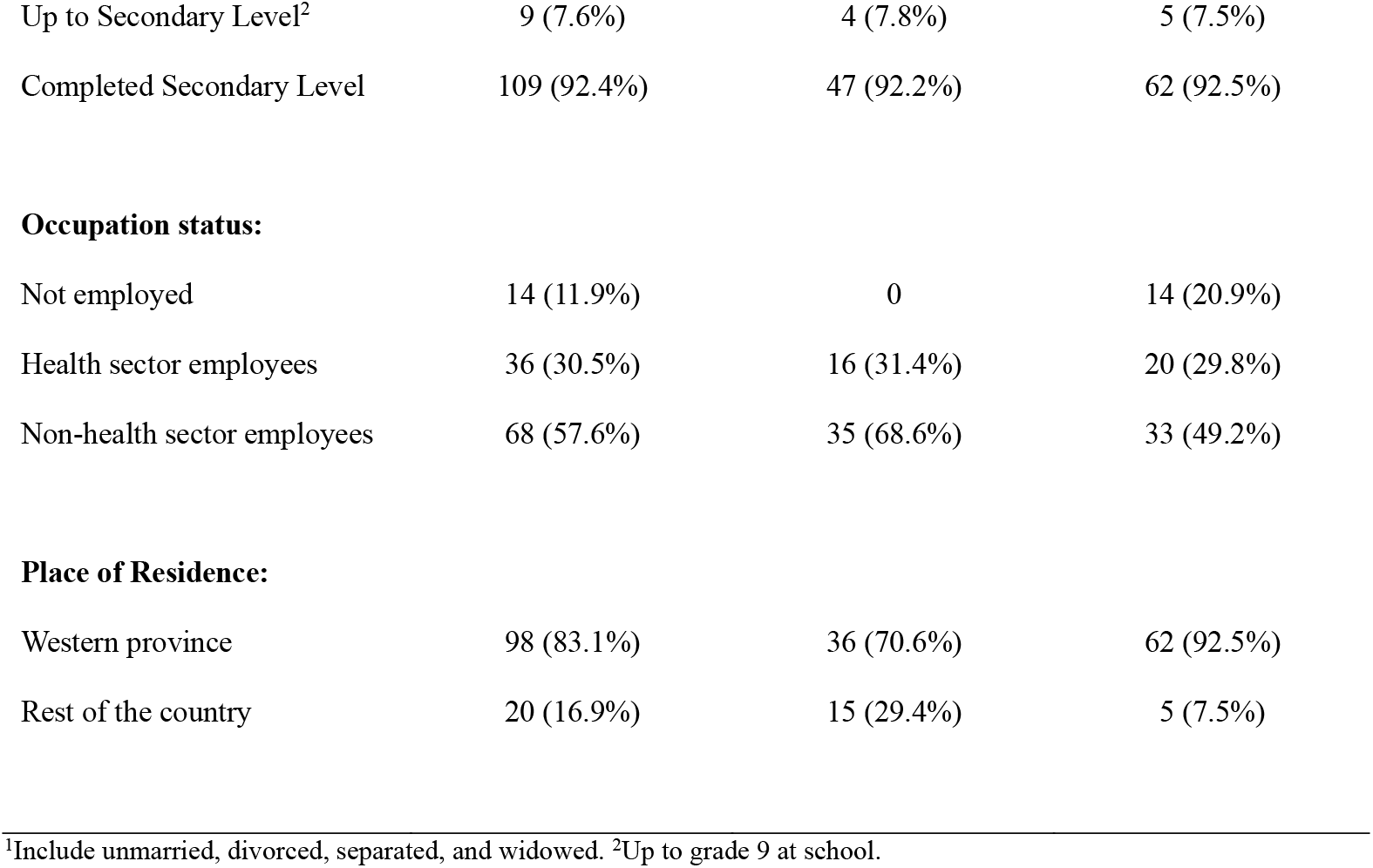
Sociodemographic status of the respondents (N=118)

| Description | Total | Males | Females |
| --- | --- | --- | --- |
|  | 118 | 51 (43.2%) | 67 (56.8%) |
| Age: |  |  |  |
| Median Age in years | 33 years | 35 years | 33 years |
| (Inter Quartile Range) | (29-38 years) | (29-39 years) | (29-36 years) |
| Civil Status: |  |  |  |
| Single<sup>1</sup> | 35 (29.7%) | 13 (25.5%) | 40 (59.7%) |
| Married | 83 (70.3%) | 38 (74.5%) | 45 (67.2%) |
| Religion: |  |  |  |
| Buddhism | 72 (61.0%) | 32 (62.7%) | 40 (59.7%) |
| Islam | 27 (22.9%) | 9 (17.6%) | 18 (26.9%) |
| Christian/ Catholic | 13 (11.0%) | 6 (11.8%) | 7 (10.4%) |
| Hinduism | 4 (3.4%) | 2 (3.9%) | 2 (2.9%) |
| Other | 2 (1.7%) | 2 (3.9%) | 0 |
| Level of Education: |  |  |  |
| Up to Secondary Level<sup>2</sup> | 9 (7.6%) | 4 (7.8%) | 5 (7.5%) |
| Completed Secondary Level | 109 (92.4%) | 47 (92.2%) | 62 (92.5%) |
| **Occupation status:** |  |  |  |
| Not employed | 14 (11.9%) | 0 | 14 (20.9%) |
| Health sector employees | 36 (30.5%) | 16 (31.4%) | 20 (29.8%) |
| Non-health sector employees | 68 (57.6%) | 35 (68.6%) | 33 (49.2%) |
| **Place of Residence:** |  |  |  |
| Western province | 98 (83.1%) | 36 (70.6%) | 62 (92.5%) |
| Rest of the country | 20 (16.9%) | 15 (29.4%) | 5 (7.5%) |

<sup>1</sup>Include unmarried, divorced, separated, and widowed. <sup>2</sup>Up to grade 9 at school.

Respondents were aware of skin cancer, asthma and mental health associated with climate change, while the knowledge on the climate impact on heart disease, diabetes, COPD and cataract of the eyes was poor (Table 2).

**Table 2.** Knowledge regarding climate change and its health effects (N=118)

|  | Good | Poor |
| --- | --- | --- |
| Knowledge on Climate Change: |  |  |
| • Heard of climate change | 116 (98.3%) | 2 (1.7%) |
| • Situations considered as components of climate change | 102 (86.4%) | 16 (13.6%) |
| • Causes for climate change | 106 (89.8%) | 12 (10.2%) |
| Knowledge on health conditions affected by climate change: |  |  |
| • Skin Cancer | 109 (92.4%) | 9 (7.6%) |
| • Asthma | 97 (82.2%) | 21 (17.8%) |
| • Mental Health | 97 (82.2%) | 21 (17.8%) |
| • Undernutrition | 78 (66.1%) | 40 (33.9%) |
| • Dengue Fever | 73 (61.9%) | 45 (38.1%) |
| • Injuries | 71 (60.2%) | 47 (39.8%) |
| • Malaria | 68 (57.6%) | 50 (42.4%) |
| • Heart Disease | 55 (46.6%) | 63 (53.4%) |
| • Cataract of the Eyes | 52 (44.1%) | 66 (55.9%) |
| • COPD | 45 (38.1%) | 73 (61.9%) |
| • Diabetes | 34 (28.8%) | 84 (71.2%) |

Most respondents had a very favourable attitude in considering climate change as a serious threat (Table 3).

**Table 3.** Attitude regarding climate change and mitigation (N=118)

|  | Good | Poor |
| --- | --- | --- |
| Attitudes on need for climate change mitigation: |  |  |
| • The weather pattern in Sri Lanka has changed from the expected predictable patterns | 102 (86.4%) | 16 (13.6%) |
| • Consider climate change as a significant issue | 115 (97.5%) | 3 (2.5%) |
| • Concerned about climate change | 110 (93.2%) | 8 (6.8%) |
| • Climate change can affect you | 115 (97.5%) | 3 (2.5%) |
| • Climate change can affect your health | 115 (97.5%) | 3 (2.5%) |
| Attitudes towards specific climate mitigation activities: |  |  |
| • Global climate change cannot be stopped or reversed, hence there is nothing we can do about it. | 93 (78.8%) | 25 (21.2%) |
| • Even a small country like Sri Lanka can do a lot of things to mitigate global climate change. | 113 (95.8%) | 5 (4.2%) |
| • Climate change is a prominent issue which our country must address as a high priority. | 116 (98.3%) | 2 (1.7%) |
| • I as an individual can make an impact to minimize climate change. | 113 (95.8%) | 5 (4.2%) |
| • Vehicle smoke emission testing is an important screening procedure for minimizing CO<sub>2</sub> emissions. | 107 (90.7%) | 11 (9.3%) |
| • Burning forests for cultivation is not justifiable. | 101 (85.6%) | 17 (14.4%) |
| • Prefer to walk over a short distance than to go in a trishaw. | 106 (89.8%) | 12 (10.2%) |
| • Cutting down trees and clearing forests is essential for economic development projects of a country. | 96 (81.4%) | 22 (18.6%) |
| • Switching off unnecessary lights and electronic devices will help reduce climate change. | 103 (87.3%) | 15 (12.7%) |
| • Chemical pesticides and chemical fertilizers are essential for a good harvest in agriculture. | 56 (47.5%) | 62 (52.5%) |
| • We need to wear protective clothing when working outdoors, when exposed to the sun. | 104 (88.1%) | 14 (11.9%) |
| • We need to ensure newly built houses have a strong roofing design to withstand hurricanes and strong winds. | 99 (83.9%) | 19 (16.1%) |
| • It is essential to plant trees along the streets in all major cities and create greener cities. | 116 (98.3%) | 2 (1.7%) |

Majority (82.2%) had a good knowledge of climate change, while most had a good attitude towards the need for climate change mitigation and climate friendly practices (Table 4).

**Table 4.**
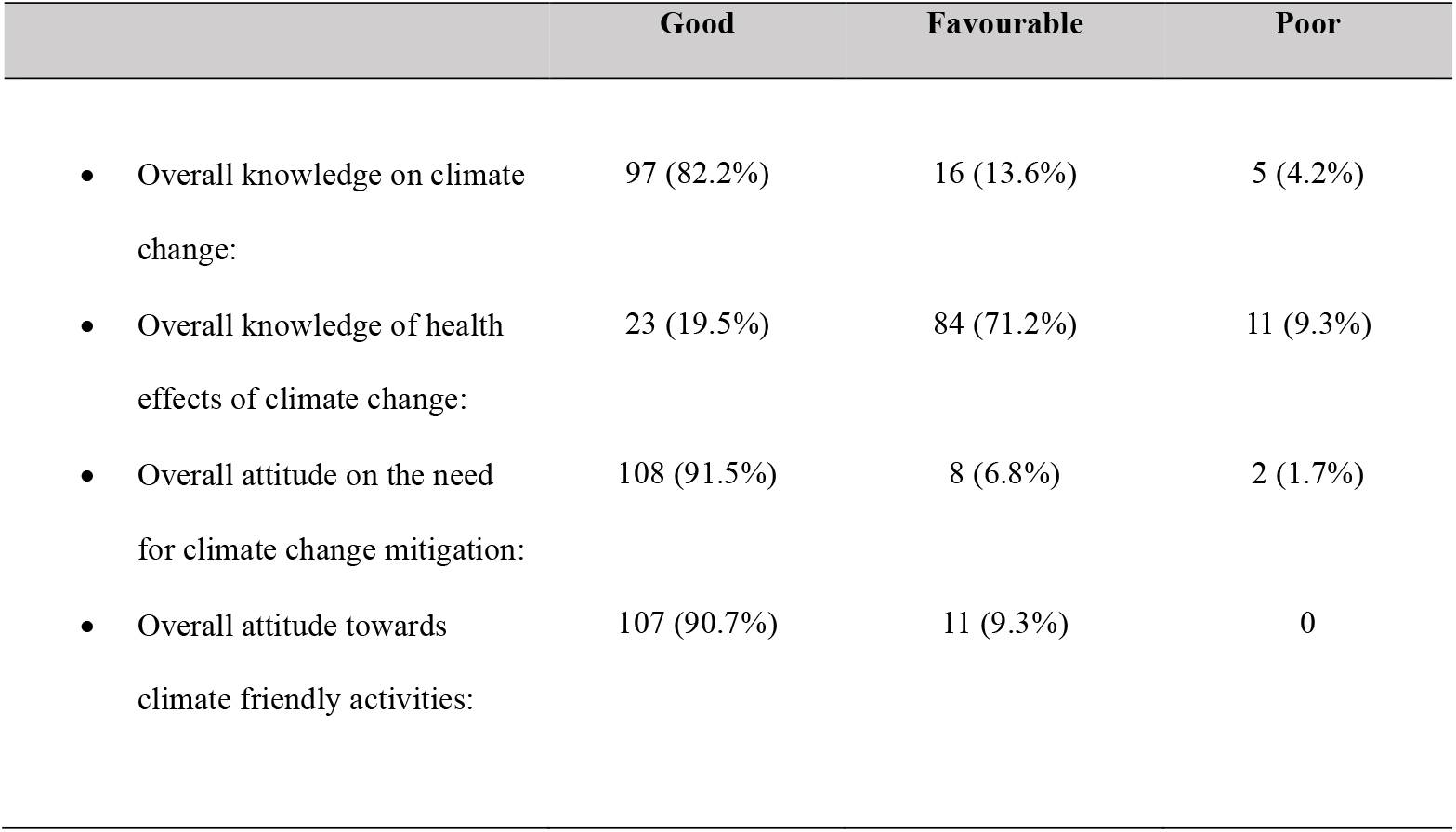
Overall knowledge and attitudes regarding climate change and impact on health (N=118)

|  | Good | Favourable | Poor |
| --- | --- | --- | --- |
| • Overall knowledge on climate change: | 97 (82.2%) | 16 (13.6%) | 5 (4.2%) |
| • Overall knowledge of health effects of climate change: | 23 (19.5%) | 84 (71.2%) | 11 (9.3%) |
| • Overall attitude on the need for climate change mitigation: | 108 (91.5%) | 8 (6.8%) | 2 (1.7%) |
| • Overall attitude towards climate friendly activities: | 107 (90.7%) | 11 (9.3%) | 0 |

Bivariate analysis of the associations between the sociodemographic factors and the knowledge or attitude regarding climate change, did not reveal any significant associations.

## Discussion

This study attempted to assess the knowledge and attitudes among the wider Sri Lankan adults, on the effects of climate change and its impact on their health. Majority of the sample were educated and employed younger adults. Majority had a good knowledge related to climate change and a varied knowledge on the health effects of climate change. Furthermore, they had a good attitude towards the seriousness of climate change and the importance of mitigating it. Most of the climate change mitigating factors were looked upon favourably but some depicted ambiguity in attitude. Overall, the knowledge and the attitudes of this sample with regards to climate change and its impact on their health and the need for its mitigation were good.

A prior preliminary survey among 54 selected staff members in a university in Sri Lanka had reported a satisfactory level of awareness on climate change and the health effects associated with it (23). A conference abstract which had assessed the impact of climate change on human health in a community division in Sri Lanka,(24) had concluded that native people are influenced by the impact of climate change, but no details were available. An abstract on children’s perceptions on health effects of climate change in Sri Lanka (25) highlight that among 104 children, there was anxiety over the issue of climate change, and they were able to identify some of the main health conditions associated with climate change. None of these available studies appear to be full text publications in the public domain, and do not report on the knowledge and attitudes of the community regarding climate change and its health impacts.

Several studies from the Caribbean region are available in this regard, especially since the Caribbean Islands are at risk of frequent adverse weather conditions.(15, 26) The study in Jamaica had reported that even though a majority of adults were aware of climate impact, about 14% were not concerned about it and continue with environmentally disruptive practices.(15) Elsewhere, a multi-Arab study among 1000 nursing students from Saudi Arabia, Egypt, Iraq and Palestine territory had reported moderate level of knowledge on the potential health impacts of climate change(27), while a study from India had reported that a majority of secondary school students had favourable knowledge of climate change.(28) A Chinese study reports of a better community awareness, on the main causes of climate change (29), and similarly, a study from America (30) had reported favourable knowledge on non-health effects of climate change among its community. In comparison, our study revealed that Sri Lankan adults had a better level of knowledge of climate change, and a much favourable attitude towards climate change mitigation. Furthermore, in our study, a vast majority correctly identified all the correct causes for climate change and had a good or favourable knowledge on the health conditions associated with climate change.

In our study, specific questions on the attitude related to the use of chemical pesticides and fertilizers, had a higher percentage of respondents with a negative attitude, and similarly, for the need for deforestation for economic development also had a higher percentage of respondents with a negative attitude. Interestingly, a fifth of the respondents were of the attitude that nothing could be done to stop or reverse climate change.

A key limitation in the current study was the small number of respondents, although it was expected that a higher number of respondents from all parts of Sri Lanka would participate due to the study being an easily accessible online survey. Also, most respondents had been from the urbanized Western province, which includes the capital Colombo and the surrounding main cities. The level of education among respondents in this current study was also quite high with most having completed secondary education. Thus, it is not possible to generalize the findings of this survey to the wider adult population of Sri Lanka. However, it helps to provide an understanding of the knowledge and attitude towards climate change and its health effects among the more educated adults of urban regions in Sri Lanka.

## Conclusions

Educated adults mainly from the urbanized western province of Sri Lanka had a good knowledge and attitude about climate change, its health impacts and the need for its mitigation. However, it is essential that more regional studies need to be conducted to understand the community awareness of the less educated people in the vast rural regions of Sri Lanka, and to implement community awareness programmes towards empowering them, since they are at a higher risk of being affected by the climate impact.

## Statements and Declarations

### Ethical considerations

Ethical clearance for the study was obtained from the Ethics review committee of the University of Colombo, Sri Lanka (EC-21-115).

### Consent to participate

Informed consent was obtained prior to the initiation of responses to the online questionnaire, and participation in the study was voluntary.

### Data Availability

Dataset is not available in the public domain but can be made available from the corresponding author upon reasonable request.

### Author contribution

IW and UM designed the study with advice from VK and AL. Study implementation and data collection were coordinated by IW, UM and TM. Manuscript was developed by IW and UM. All authors reviewed and commented on the manuscript and refined it appropriately. All authors reviewed and approved the final version of the manuscript for submission.

